# Genetic Regulation of the Metabolome Differs by Sex, Alzheimer’s Disease Stage, and Plasma Biomarker Status

**DOI:** 10.1101/2025.02.26.25322932

**Authors:** Jaclyn M. Eissman, Min Qiao, Vrinda Kalia, Marielba Zerlin-Esteves, Dolly Reyes-Dumeyer, Angel Piriz, Saurabh Dubey, Renu Nandakumar, Annie J. Lee, Rafael A. Lantigua, Martin Medrano, Diones Rivera Mejia, Lawrence S. Honig, Clifton L. Dalgard, Gary W Miller, Richard Mayeux, Badri N. Vardarajan

**Author notes:** Address Correspondence to: Badri N. Vardarajan, PhD, Taub Institute, G.H. Sergievsky Center, Columbia University, New York, NY 10032.

## Abstract

We investigated genetic regulators of circulating plasma metabolites to identify pathways underlying biochemical changes in clinical and biomarker-assisted diagnosis of Alzheimer’s disease (AD). We computed metabolite quantitative trait loci by using whole genome sequencing and small molecule plasma metabolites from 229 older adults with clinical AD and 322 age-matched healthy controls. Unbiased associations between 6,881 metabolites and 332,772 common genetic variants were tested, adjusted for age, sex, and both metabolomic and genomic principal components. We identified 72 novel and known SNP-metabolite associations spanning 66 genes and 12 metabolite classes, including *PYROXD2* and N6-methyllysine, *FAAH* and myristoylglycine, as well as *FADS2* and arachidonic acid. In addition, we found differences in genetic regulation of metabolites among individuals with clinically defined AD compared to AD defined by a published plasma P-tau181 level cut-off. We also found more SNP-metabolite associations among males compared to females. In summary, we identified sex- and disease-specific genetic regulators of plasma metabolites and unique biological mechanisms of genetic perturbations in AD.

## Introduction

Metabolic changes are part of Alzheimer’s disease (AD) pathogenesis, beginning very early in disease^1^ with a decline in glucose metabolism in the brain.^2^ Throughout disease, other metabolic shifts include changes in lipids, phosphatidylcholines, ceramides, and lysophosphatidylcholines, as well as in bile acid metabolism^3^, methylhistidine metabolism, and fatty acid metabolism.^3,4^ Prior studies show that metabolic signatures can accurately discriminate healthy controls, individuals with mild cognitive impairment (MCI), and those with AD, especially between the latter two groups.^5–8^

Twin studies show that endogenous metabolic features may be heritable, estimating individual locus contributions at a median of 6.9% and a maximum of 62%,^9^ and likewise SNP-based estimates find a strong median SNP-based heritability of 19.7%.^10^ Multiple quantitative trait loci (QTL) studies have found robust genetic loci associated with the metabolome.^10–15^ However, most of these studies have not focused on aging- and AD-specific genetic regulation, and have been predominantly in non-Hispanic whites, limiting the generalizability of findings to more diverse populations.^15^ It is important to explore genetic regulation of the metabolome in diverse groups, as AD risk differs by race and ethnicity^16^ and metabolic changes in those with AD likewise differ by race and ethnicity.^17^

In addition to racial and ethnic differences, sex differences are apparent in AD risk and pathogenesis, with robust sex differences in prevalence,^16^ lifetime risk,^16^ as well as neuropathologic burden and its relation to clinical AD presentation.^18,19^ Evidence also suggests sex differences in the metabolome^20^ and in metabolic dysregulation in AD.^3,7,21^ Arnold and colleagues^3^ identified sex differences in associations of metabolites and AD biomarkers, which included acylcarnitines and amino acids showing sex-specific associations with CSF P-tau and FDG-PET.^3^ Furthermore, modules containing metabolites such as phosphatidylcholines, acylcarnitines, lipids, amino acids, and sphingomyelins showed sex-specific associations with AD brain endophenotypes.^1^ Genome wide association studies (GWAS) studies have identified sex-specific loci associated with AD endophentoypes,^22–26^ but the intersection of genetic regulation and the metabolome in AD has not been fully elucidated.

The goal of this study is to expand our understanding of genetic regulation of the metabolome in AD and to elucidate the impact of sex and AD pathology on this relationship. We conducted a genome- and metabolome-wide QTL analysis in a Caribbean Hispanic cohort of aging and AD. We conducted metabolome QTL analyses in the full cohort, by clinical and P-tau181-assisted diagnosis subgroups, and by biological sex. This study adds to the current literature by clarifying the relationship of genetic relation of the metabolome in a diverse sample, by sex, and by clinically and biomarker-assisted AD diagnosis.

## Results

### Study Participants

After both genetic and metabolomic data quality control, 551 Estudio Familiar de Influencia Genetica en Alzheimer (EFIGA; **Table 1**) study participants were included in this analysis, with 229 clinical AD cases and 322 age-matched healthy clinical controls. The full sample consisted of 153 men (27.77%) and 398 women (72.23%). The average age was 71.08 (+/-7.89) years among the full sample. Furthermore, 353 participants (64.07%) were *APOE* ε4 non-carriers and 195 (35.39%) were *APOE* ε4 carriers (and 3 [0.54%] were missing *APOE* information). Additionally, 384 participants (69.69%) were classified as biomarker-negative controls (P-tau181<2.63) and 163 participants (29.58%) were classified as biomarker-supported AD (P-tau181>/=2.63; and 4 [0.73%] were missing biomarker status).^27^

**Table 1.** Participant Characteristics of Individuals Included in this Study from the EFIGA Cohort.

|  | **Healthy Controls**<br>**(N=322)** | **Clinical AD Cases**<br>**(N=229)** | **Full Sample**<br>**(N=551)** |
| --- | --- | --- | --- |
| **Age at last visit** |  |  |  |
| Years, Mean (SD) | 68.71 (+/- 7.02) | 74.38 (+/- 7.86) | 71.08 (+/- 7.89) |
| **Sex** |  |  |  |
| Males, N(%) | 81 (25.16%) | 72 (31.44%) | 153 (27.77%) |
| Females, N(%) | 241 (74.84%) | 157 (68.56%) | 398 (72.23%) |
| **Plasma P-tau181 level** |  |  |  |
| <2.63 (-), N(%) | 247 (76.71%) | 137 (59.83%) | 384 (69.69%) |
| ≥2.63 (+), N(%) | 72 (22.36%) | 91 (39.74%) | 163 (29.58%) |
| Missing, N(%) | 3 (0.93%) | 1 (0.44%) | 4 (0.73%) |
| **APOE ε4 allele** |  |  |  |
| Non-carrier, N(%) | 221 (68.63%) | 132 (57.64%) | 353 (64.07%) |
| Carrier (1+ ε4), N(%) | 99 (30.75%) | 96 (41.92%) | 195 (35.39%) |
| Missing, N(%) | 2 (0.62%) | 1 (0.44%) | 3 (0.54%) |

### Genome-Wide Metabolite Quantitative Trait Loci Analysis

Metabolite QTL (Met-QTL) analyses were performed separately for C18- and HILIC+ columns, adjusting for a genome-wide false-discovery rate of (FDR<0.05) among each set of tests. Overall, we identified 72 Met-QTLs that survived both adjustment for multiple comparisons (FDR<0.05) and post-hoc filtering criteria (**Figure 1A; Supplementary Table 1**). These 72 QTLs spanned 66 unique genes and 12 unique metabolite classes. Four significant Met-QTL pairs were also observed in previously published QTL studies. The strongest association, *PYROXD2* and N6-methyllysine (amino acid), was validated in published data from three QTL studies,^10,11,15^ whereby each study also identified *PYROXD2* and N6-methyllysine. Similarly, one more study^12^ previously identified *PYROXD2* and observed an association also with a lysine derivative, N-methylpipecolate. We additionally validated the association between *FAAH* and myristoylglycine (fatty amide, N-acyl amine) in data from three previously published QTL studies^10,12,13^ all of whom identified associations between *FAAH* and acylglycines (N-acyl amines). We found an association with *PDXDC1* and trans-2-Dodecenoylcarnitine, an acylcarnitine, and previous work^10^ likewise identified an association between *PDXDC1* and an acylcarnitine. Interestingly *FADS2* regulates lysophosphatidylcholines (lysoPC) that bind to or interact with arachidonic acid. The association between *FADS2* and arachidonic acid was observed in both our study and a recent, published QTL study,^12^ in another study between *FADS2* and both a related molecule to arachidonic acid and multiple phospholipids,^13^ between *FADS2* and lysophosphatidylcholines in another study,^14^ and finally between *FADS2* and a glycerophosphocholine, 1-palmitoyl-2-dihomo-linolenoyl-GPC (16:0/20:3n3 or 6) in a very recent QTL study.^15^ Additionally, from the candidate AD Met-QTL analysis, we identified 35 Met-QTL associations passing the *a priori* significance threshold of p<10^-4^ (**Supplementary Table 12**).

**Figure 1.**
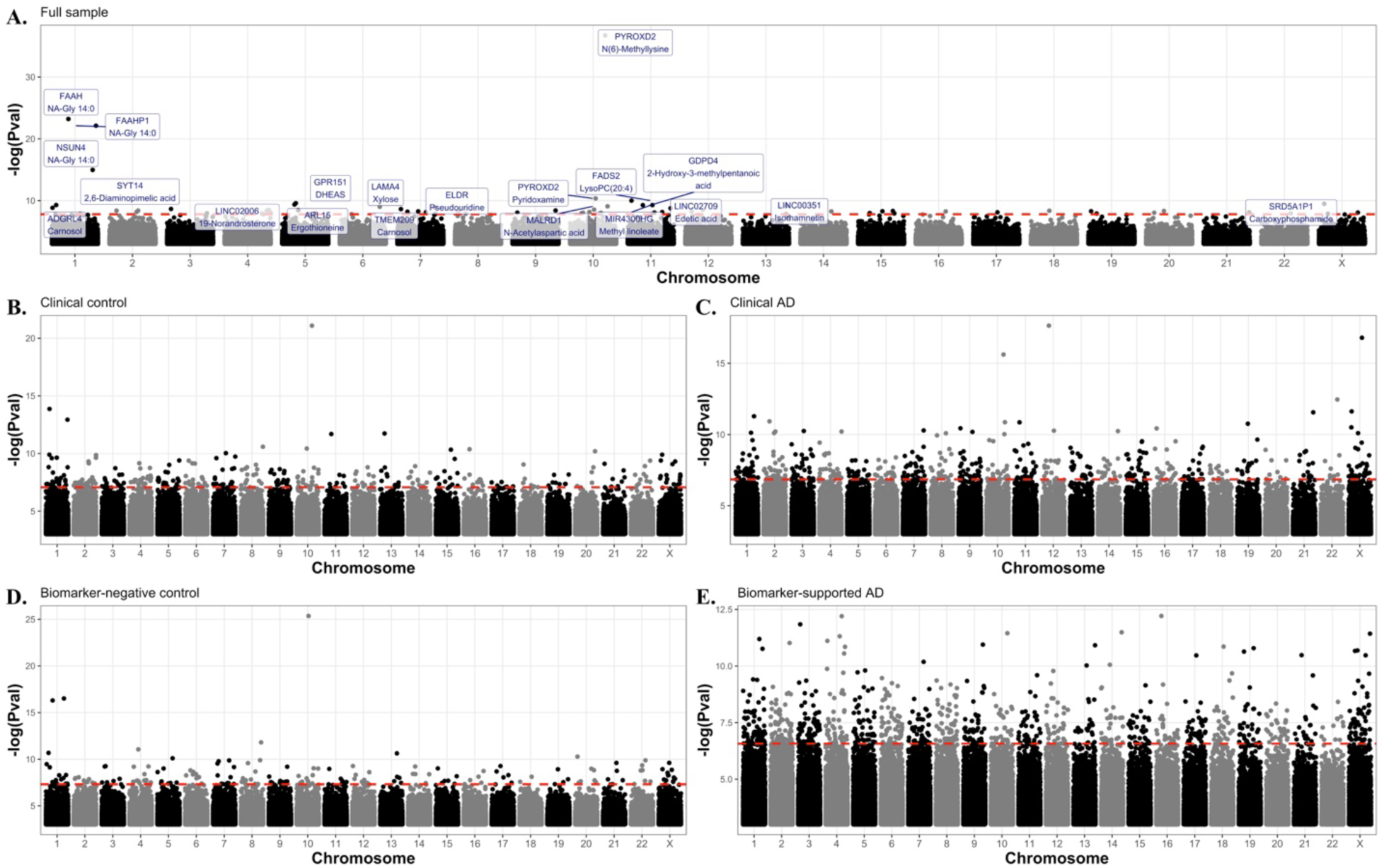
Genome and Metabolome-Wide QTL Study in Full Sample and by Diagnosis. Manhattan plots depicting metabolome-wide QTL study results among the full sample (A), among individuals who are clinically healthy (B), those with clinical AD (C), individuals who are biomarker-negative controls (D), and individuals with biomarker-supported AD (E). The red dashed lines depict the significance threshold, determined based on a genome-wide false-discovery rate threshold (FDR<0.05). Among the full sample (A), the top 20 QTLs are annotated in blue.

### Genome-Wide Metabolite Quantitative Trait Loci Subgroup Analysis

We identified Met-QTLs in participants stratified by clinical diagnosis of cognitively unimpaired or clinical AD (**Figure 1B-C**; **Figure 3A-B; Supplementary Tables 2-3**). Among clinical controls, we identified 308 Met-QTLs surviving adjustment for multiple comparisons (FDR<0.05) and post-hoc filtering criteria, with 274 non-overlapping QTLs with AD QTLs. Among those with clinical AD, we identified 507 Met-QTLs surviving adjustment for multiple comparisons (FDR<0.05) and post-hoc filtering criteria, of which 463 were not observed in healthy participants. Next, we stratified the sample based on AD diagnosis defined by P-tau181 levels (**Figure 1D-E**; **Figure 3A-B; Supplementary Tables 4-5**). Among biomarker-negative controls, we identified 190 Met-QTLs surviving adjustment for multiple comparisons (FDR<0.05) and post-hoc filtering criteria, with 168 non-overlapping QTLs with biomarker-supported AD. Among those with biomarker-supported AD, we identified 866 Met-QTLs surviving adjustment for multiple comparisons (FDR<0.05) and post-hoc filtering criteria, with 799 non-overlapping QTLs with biomarker-negative controls. Notably, there were significant QTLs that were present among both clinical AD cases and individuals with biomarker-supported AD, but not among either control group. One noticeable difference between both healthy control-strata as compared to clinical AD and biomarker-supported AD was that we found nearly one-third more QTLs associated with fatty acids among clinical AD compared to healthy controls and greater than 7-fold more QTLs associated with fatty acids among biomarker-supported AD cases as compared to biomarker-negative controls. For example, we observed SNP associations with Eicosapentaenoic fatty acid in clinical AD and biomarker-supported AD groups but not in either healthy-control strata.

### Sex-Stratified Genome-Wide Metabolite Quantitative Trait Analysis

We performed sex-stratified Met-QTL analyses in males and females, clinically healthy and with clinical AD (**Figure 2**; **Figure 3C-D; Supplementary Tables 6-11**). Among all males (**Figure 2A; Supplementary Table 6**), 687 Met-QTLs survived adjustment for multiple comparisons (FDR<0.05) and post-hoc filtering criteria, with 632 non-overlapping QTLs with females. Among all females (**Figure 2D; Supplementary Table 9**), 137 Met-QTLs survived adjustment for multiple comparisons (FDR<0.05) and post-hoc filtering criteria, with 120 non-overlapping QTLs with males. Types of QTLs that tended to differ between sexes included those associated with fatty acids and glycerophospholipids, both of which were more prevalent among males, and moreover the types of fatty acids associated also differed between sexes. Further, there were significant differences between the QTLs identified in male and female clinically healthy participants and males and females with clinically diagnosed AD (**Figure 2B-C,E-F, Supplementary Tables 7-8 & 10-11**).

**Figure 2.**
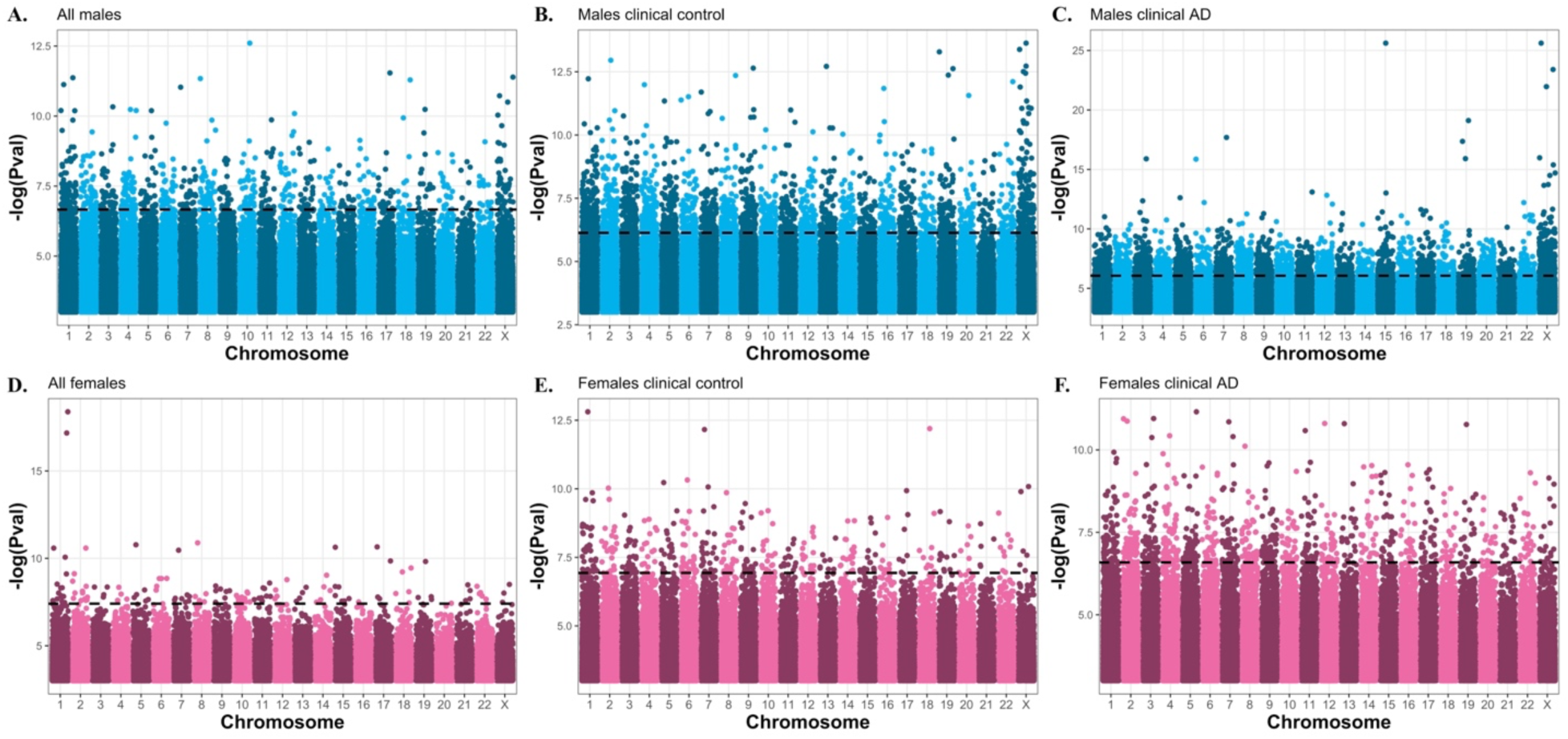
Sex-Specific Genome and Metabolome-Wide QTL Study in Full Sample and by Diagnosis Strata. Manhattan plots depicting metabolome-wide QTL study among males (blue) and females (pink). The top row shows results in the full sample of males (A), males who are clinical controls (B), and males who have clinical AD (C). The bottom row shows results in the full sample of females (D), females who are clinical controls (E), and females with clinical AD (F). The black dashed lines depict the significance threshold, determined based on a genome-wide false-discovery rate threshold (FDR<0.05).

## Discussion

We performed a genome- and metabolome-wide association study in a cohort of older adults with and without AD. We identified 72 robust QTLs across 66 genes spanning the genome, and these QTLs were associated with metabolites across 12 unique classes. Notably, we validated some of these gene-metabolite pairs in previous QTL studies,^10–15^ and our study clarified that genetic regulation of these metabolites may be implicated in AD pathogenesis. Sex- and diagnosis-specific analyses clarified that most QTLs in the metabolome do not appear to be shared across sexes and diagnostic subgroups (**Figure 3**), providing evidence that genetic regulation of metabolic changes with age and in AD possibly differ by sex and by disease stage.

**Figure 3.**
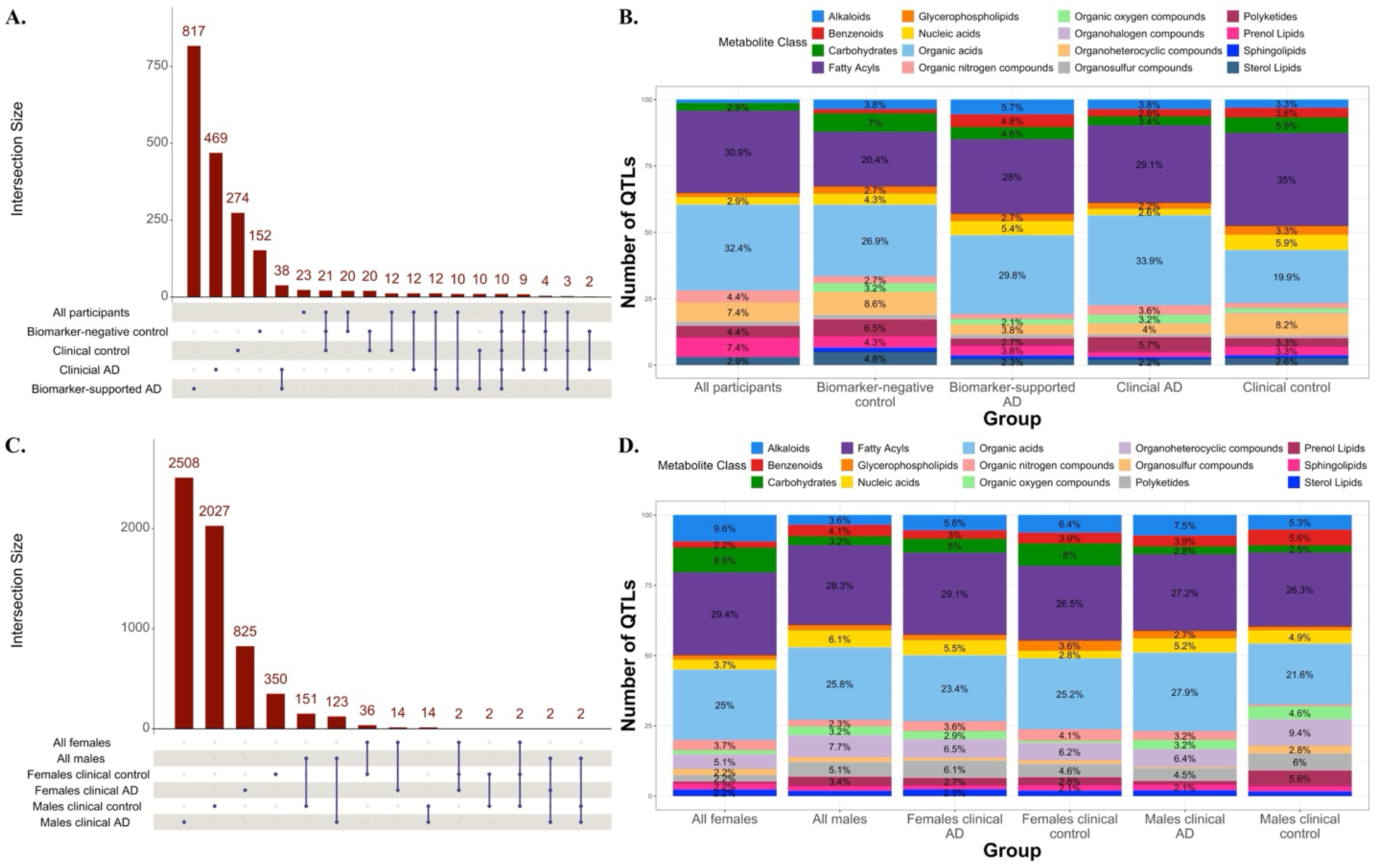
Unique and Shared Metabolite-QTLs Across Sexes and Diagnosis Subgroups. Upset plots (A) and (C) illustrate the overlapping and non-overlapping QTLs among diagnosis subgroups (A) and among sexes (C). Blue dots connected by lines under each set of bars show which subgroups share QTLs, whereas a blue dot with no line under a bar indicates those QTLs are not shared across any subgroups. (B) and (D) show the percentage of QTLs that belong to each metabolite class.

Validated Met-QTL associations include, *PYROXD2* and N6-methyllysine,^10–12,15^ *FAAH* and myristoylglycine,^10,12,13^ *FADS2* and arachidonic acid (LysoPC(20:4)),^12–15^ and *PDXDC1* and an acylcarnitine (trans-2-Dodecenoylcarnitine).^10^ *PYROXD2* has been identified in multiple QTLs studies in association with N6-methyllysine or methyl-L-lysine derivatives.^28^ Lysine metabolism plays a reported role in AD pathology, including that methylation of lysine residues is a post-translational modifier of tau in neurofibrillary lesions,^29,30^ and may harbor some protection against tau pathological aggregation.^30^ One study further demonstrates that lysine metabolic changes can differentiate cognitively unimpaired individuals from those with MCI or AD.^5^ Interestingly, in our study the *PYROXD2*/N6-methyllysine pair was present in both controls and AD groups, as well as among biomarker-negative controls and biomarker-supported AD groups, but showed more significant associations among both healthy control strata (**Supplementary Tables 1-5**).

Validated association, *FAAH* and myristoylglycine, was observed among clinical and biomarker-negative controls as compared to those with AD and biomarker-supported AD (**Supplementary Tables 1-5**). N-acyl glycines, such as myristoylglycine, are upregulated in AD.^5^ Furthermore, fatty acid amide hydrolase (*FAAH*) enzyme inhibitors play a known role in AD.^31^ *FAAH* is found in the brain and is part of the endocannabinoid pathway, a well-established pathway in AD etiology, whereby endocannabinoid levels correlate with AD biomarkers, especially Aβ pathology and memory performance.^31^ Interestingly, *FAAH* and myristoylglycine was one of the strongest associations among females, and also is significant among females who were clinical controls or had clinical AD (**Supplementary Table 9-11**). This reveals that the main effects association was likely driven by females, suggestive that this metabolic change may be important in the clinical manifestation of AD among females more than males. It also reemphasizes the importance of sex-specific analysis for not only identifying novel genetic loci but for uncovering information about associations identified in a main effects analysis.

*FADS2* and arachidonic acid (LysoPC(20:4)), a top, validated association in this study, does not appear associated with clinical or biomarker-negative controls nor clinical AD strata (**Supplementary Tables 2-4**). However, this association does appear in the biomarker-supported AD strata (**Supplementary Table 5**). The *FAD* gene cluster, which includes *FADS2*, is associated with cognition, whereby a Mendelian randomization colocalization analysis illustrates that *FADS1* and *FADS2* expression appear to have a causal effect on cognition.^32^ Furthermore, genetic variation in the *FAD* gene cluster is associated with arachidonic acid levels, and arachidonic acid interacts with Aβ40 and Aβ42 pathology.^33^ Since *FADS2*/arachidonic acid was a top association in the main analysis and in biomarker-supported AD only, and arachidonic acid plays a role in AD pathology, this may mean that genetic regulation of the *FAD* gene cluster and arachidonic acid play an important role once AD pathology is present, but not before.

*PDXDC1* and trans-2-Dodecenoylcarnitine, an acylcarnitine, was a validated association in the main effects analysis and then appeared only in clinical controls (**Supplementary Tables 1-5**). Acylcarnitine levels in plasma are increased in individuals with AD,^4^ and furthermore these metabolites show a pattern of consistent decrease from preclinical to clinical AD and may be able to identify AD converters before onset of disease.^4,34^ One key feature of acylcarnitines is that they tend to show sex-specific effects,^1,3^ and notably in our sex-stratified analyses, both among males and females, we identified QTLs associated with various acylcarnitines (**Supplementary Table 6-11**). Taken together, future studies should continue to investigate the sex-and disease-specific role of individual acylcarnitines in AD etiology.

Our analysis shed light on sex-biased genetic regulation of the metabolome. Sex differences in metabolic changes in AD^3,7,21^ are well understood, but the question remains if shared genetic variation or if differing genetic variation mainly contributes to these observed phenotypic differences. This analysis provides evidence that most genetic variation relating to the metabolome in AD differs between sexes, as shown in **Figure 3**, very few QTLs are shared between sexes. Importantly, the sex-specific QTLs are not due to metabolite level differences by sex, as we filtered out any Met-QTLs whereby the metabolite levels showed significant differences between sexes. In addition, the sex-specific QTLs were spread throughout the genome, spanning beyond sex chromosome complement differences. One possible contribution to the observed sex differences is sex hormones, as a sex-specific relationship already exists between sex hormones, aging, and AD.^35^ Future studies should further investigate the sex-specific crosstalk between the genome, the metabolome, and sex hormones in aging and AD.

Strengths of this study include the untargeted metabolomics approach which both allows for more metabolites to be included and is a less biased approach as compared to targeted metabolomics. Additional strengths include the diverse sample, as most large Met-QTL studies have only been performed in non-Hispanic white individuals. The inclusion of sex-specific models, including the X-chromosome, allowed for a more complete understanding of the association of the genome and metabolome in each sex. Weaknesses of this study include the sample size, as we had 551 individuals in this study and recent QTL studies had sample sizes in the thousands. Our sex-stratified and diagnosis-stratified models also had imbalances, for example, we had fewer males compared to females, which could have partially influenced this analysis. Furthermore, we did not report QTL associations with metabolites that were not properly annotated and of an unknown class, but these metabolites do have meaning and should be investigated in future studies. Lastly in this study, we covaried for *APOE* ε4 carrier status to investigate the genetic regulation of the metabolome above and beyond the well-characterized *APOE* locus, but we do know that *APOE* is involved in lipid metabolism and shows sex differences, making it a key player in the sex-specific relationship of the genome and metabolome.

Overall, this analysis identified both novel and validated genetic regulators of the metabolome in aging and AD in a Hispanic cohort. We provide hundreds of novel sex-specific and disease-specific Met-QTL findings that have not previously been the focus of most QTL studies to date, especially in the context of AD. Future studies should continue to investigate the relationship of the genome and the metabolome in AD through a precision medicine lens to continue to better understand the totality of the genetic contribution to AD at the molecular level.

## Methods

### Study Participants

This study included participants from the Estudio Familiar de Influencia Genetica en Alzheimer study (EFIGA).^36^ Recruitment for EFIGA began in 1998, following participants every 2 years, enrolling individuals of Caribbean Hispanic ancestry from the Dominican Republic and the Washington Heights area of New York. All study participants had late-onset AD (LOAD) or a family history, and were given a standardized evaluation, including a neurological test battery, structured medical and neurological exams, and a depression assessment.^37,38^ LOAD diagnoses were determined from the NINCDS-ADRDA criteria^39,40^ for probable or possible LOAD^41^, and the Clinical Dementia Rating^42–44^ was included to determined disease severity. Hixson and Vernier^45^ modified criteria^46^ and Taqman genotyping were leveraged to determine each participant’s *APOE* genotype.

### Whole Genome Sequencing Data Collection

Whole genome sequencing (WGS) was performed at the Uniformed Services University Health Sciences leveraging an Illumina PCR-free library protocol, sequencing the data on the Illumina NovaSeq platform. WGS was generated among individuals with clinically diagnosed AD and age-matched healthy controls using DNA extracted from PAXgene tubes at a mean coverage of 30×. Analysis of WGS data was performed with an automated pipeline which is in line with recommendations from the Centers for Common Disease Genomics (CCDG) and the Trans-Omics for Precision Medicine (TOPMed) platforms.^47^ Reads were aligned to human reference hs38DH with BWA-MEM v0.7.15, and variant calling was conducted according to recommendations from the Genome Analysis Toolkit.

### Whole Genome Sequencing Quality Control

Using BCFtools v1.19, we split multi-allelic variants, aligned indels, retained variants that pass all filters, and set missing genotypes to the reference genotype. VCF files were converted to PLINK (v2.0 and v1.9) binary file sets retaining only biallelic SNPs, filtering for mean depth >10 and genotype quality >20. On the binary file sets, we performed variant-level filtering, including filtering out variants with a missing genotyping rate >5% and retaining common polymorphisms at a minor allele frequency (MAF) of >5%. Then we performed sample-level filtering, including filtering out samples with >1% sample missingness, and dropping duplicate samples. We conducted identify-by-descent relatedness calculations, dropping both samples in a pair if a pi-hat estimate was >0.9, and one sample of a pair if a pi-hat estimate was between 0.25 and 0.9. Specific X-chromosome processing included removing the pseudo-autosomal region, sex check and sex imputation (for missing sex), as well as a differential missingness test between sexes (p<10^-7^). A Hardy-Weinberg Equilibrium (HWE) exact test was conducted in all samples (p<10^-6^) and among females for the X-chromosome, filtering male samples accordingly. Additionally, to ensure only common variants were retained, we compared variant frequencies to gnomAD, keeping variants with >5% gnomAD frequency. Finally, we performed a principal component analysis (PCA; with PC-AiR) to assess genetic ancestry and cryptic relatedness, removing sample outliers with an iterative outlier removal procedure. The final, cleaned genetic data consisted of 619 samples (452 females and 167 males) and 6,665,147 variants.

### Plasma Metabolomics Data Generation

The protocol for metabolite data generation was previously described.^48^ Plasma was collected by venipuncture in K2EDTA tubes, and by 2 hours of collection was centrifugated, prepared, and store at −80°C.^27,48^ Metabolites were extracted with acetonitrile, and injected in triplicate into two chromatographic columns: a hydrophilic interaction column under positive ionization (HILIC+)^49^ and a C18 column under negative ionization (C18-),^50^ which resulted in 3 technical replicates per sample per column. Columns were coupled to a Thermo Orbitrap HFX Q-Exactive mass spectrometer and scanned for 85 – 1250 kDa molecules. To process the metabolites, feature detection and peak alignment were performed with apLCMS^51^ and xMSanalyzer^52^ software. Feature tables were produced that included mass-to-charge ratio, retention time, and median summarized abundance/intensity of each ion (i.e., metabolic feature) for each sample. Then an empirical Bayesian framework batch correction was implemented with ComBat.^53^ Metabolic features were retained if present in 70% or greater of samples, resulting in 3,253 features and 3,628 features from the HILIC+ and C18-columns, respectively.^48^ If a feature had zero-intensity, it was deemed to be below the detection limit and for each of these features a ½ minimum intensity for each observed metabolic feature was leveraged to impute the value. All features were log-transformed, quantile normalized, and auto scaled.^48^

### Metabolite Annotation

Processed metabolic features were annotated leveraging the Human Metabolome Database (HMDB) and a multi-stage clustering algorithm from the R package, xMSannotator (v.1.3.2).^48,52^ Annotation was implemented for metabolic features in order to get pathway associations, intensity profiles, retention time, mass defect, and isotope/adduct patterns. A confidence level of 1 to 5 was assigned with each annotation, which was determined based on a previous published protocol^54^ and levels 1-3 (where 1 = most confident) were used in this analysis. To assign a singular annotation when multiple annotations matched one feature, a protocol was followed for annotation that included the following rules: First, annotations were chosen based on the highest confidence score. Second, if scores were similar then the lowest difference between expected vs. observed mass was chosen. If these strategies did not resolve annotations, then a feature was labeled as having multiple matches or unknown.

### Metabolome Quantitative Trait Loci Analyses

Prior to performing the metabolome quantitative trait loci (Met-QTL) analyses, we retained individuals who had both WGS data and metabolomics data, which resulted in 551 individuals. To ensure robust data quality amongst this specific sample, we performed additional quality control steps, including a specific MAF filter (>5%) and HWE test (p<10^-6^) among the N=551. Due to the high multiple testing burden and non-independence between variants (linkage disequilibrium - LD), we LD-pruned the genetic data prior to analysis, resulting in 332,772 variants carried forward to analysis. All QTL analyses were performed with the MatrixEQTL R package (v. 2.3)^55^ applying the linear association model and a genome-wide false discovery rate (FDR) adjustment, with *a priori* significance set at FDR<0.05. QTL models included each metabolite as the outcome and age, sex, a binary diagnosis variable (cognitively unimpaired or AD), and *APOE* ε4 carrier status (binary variable) as covariates. Additional covariates included the first three genetic ancestry principal components and the first three principal components of the metabolomics data. Met-QTL subgroup analyses included 1) sex-specific models, stratifying by sex (male or female), and 2) diagnosis-specific models, stratifying by binarized clinical diagnosis (cognitively unimpaired or AD). Exploratory models stratified by P-tau181 status, using a previously defined cut-off^27^ creating two groups: biomarker-negative control and biomarker-supported AD.

### Candidate Met-QTL Analysis of Published Alzheimer’s Disease GWAS Loci

We tested whether previously published AD risk and protective loci were associated with metabolites, potentially elucidating biological mechanisms to these loci by understanding their influence on the metabolome. To conduct these tests, we compiled AD GWAS loci from the Bellenguez et al^56^ manuscript. In total, we tested 76 AD loci to clarify their relationships to the metabolome in a candidate analysis to reduce multiple testing burden, and we set an *a priori* significance threshold at p<10^-4^.

### Prioritization of Met-QTLs

We applied a four-step filtering procedure to prioritize Met-QTLs. We retained QTLs with both a p-value surviving genome-wide FDR correction and that mapped to a metabolite with an annotation confidence level of 1-3.^54^ Additionally, we mapped metabolites to previously known classes, and if an annotation was missing, we filtered out the QTL. We also retained QTLs that mapped to a known gene (ANNOVAR, 2020-06-07 release).^57^ If variants were mapped to multiple genes, we selected the gene that was in closest proximity to the variant location, or if ambiguous, we randomly selected a listed gene. If a variant mapped to genes greater than 1Mb away, we set the gene as missing. For sex-specific analyses, we performed one more filtering step, removing Met-QTL pairs whereby the metabolite levels were significantly different between sexes. This retained only QTLs where the genetic regulation of metabolites differed by sex and not the levels of the metabolite itself. Group differences were determined by comparing mean levels between males and females through Welch’s two-sample t-tests (in R), whereby a metabolite was considered as significantly different between sexes if the t-test had a p<0.05.

### Validation of Significant QTLs from Met-QTL Analyses

In an effort to validate our main effects results, we surveyed large blood, plasma, CSF, or brain metabolite QTL studies including Chen et al.,^10^ Yin et al.,^11^ Hysi et al.,^12^ Long et al,^13^ Lotta et al.,^14^ and Wang et al.^15^ Our study mapped QTLs to genes, and thus we compared our set of prioritized genes from the significant QTLs to that of each study above. If a gene matched and the associated metabolite was identical or of the same metabolite class as in our study, we considered it as evidence for validation.

## Data availability

All results are included in the main text or the supplementary materials of the manuscript. The raw whole genome sequencing, metabolomics and biomarker data will be shared with qualified investigators using the request form available here: https://cumc.co1.qualtrics.com/jfe/form/SV_dmck0uV3A91pmzb. The WGS data is also available via the Alzheimer’s Disease Sequencing Project: https://dss.niagads.org/datasets/ng00067/.

## Code availability

Code from this study is made available on GitHub: https://github.com/jaclyn-eissman/Metabolome-Wide-QTL-Analysis.

## Acknowledgements

EFIGA study is supported by NIA grants R56AG063908, R01AG067501 and RF1AG015473. We acknowledge the services of CEDIMAT for collaborating with sample collection and processing in the EFIGA cohort. The metabolomics core that generated the metabolomics data for the project is supported by the National Center for Advancing Translational Sciences grant-5UL1TR001873. Additionally, we thank Drs. Carlos Cruchaga and Postdocs, as well as Ph.D. students for their valuable input in validating the results from the Met-QTL analyses in this dataset.

## Author information

Author contributions Conceptualization: JME, BNV

Methodology: JME, BNV, CLD, RN, VK, GWM

Participant enrolment and Sample Collection: DRD, MM, DRM, RAL, LSH, RPM

Data Generation: MQ, VK, AP, MZE, RN, SD, CLD, AJL, LSH, GWM, RPM, BNV

Data analysis and interpretation: JME, BNV, VK

Funding acquisition: RPM, GWM, BNV

Project administration: DRD, RAL, RPM

Supervision: BNV

Writing: original draft: JME, BNV

Writing: reviewing & editing: all authors

